# Application of hybrid failure mode effect analysis based on fuzzy AHP and fuzzy TOPSIS in quality risk assessment of fixed-dose artemether-lumefantrine tablets

**DOI:** 10.1101/2024.01.26.24301832

**Authors:** Salim Ilyasu, Sani Malami, Garba Mohammed Khalid, Umar Sharif Abdussalam, Abubakar Magaji Dawud, Saratu Mohammed, Al-Kasim Rabiu Falalu, Mustapha Fatima Zaharadeen, Farouq Idris Sani, Abdussalam Yayo Manu, Ibrahim Adamu Yakasai, Basheer Abba-Zubair Chedi, Aminu Sahalu Bello, Adam Bashir Ibrahim, Saadatu Julde, Maryam Shehu Idris, Lawal Alhassan Bichi

## Abstract

**Background:** A hybrid Failure Mode Effect Analysis (FMEA) based on fuzzy logic and multicriteria decision analytics was applied as a post-marketing surveillance tool for troubleshooting potential quality problems of Artemisinin Combination Therapies (ACTs) as a means of aggregating subjective opinions of quality experts in providing vital information for continuous management and improvement of safety and quality standards.

**Research design and methods:** An FMEA team of five cross-functional quality experts implemented a hybrid fuzzy FMEA model based on Analytical Hierarchy Process (AHP) and Technique for Order of Preference by Similarity to Ideal Solution (TOPSIS) to solve multicriteria decision problems in quality risk analysis of historic quality control data of ACTs.

**Results:** Expert opinions were captured as triangular fuzzy numbers to represent the linguistic scoring of risk-determining variables for fifteen failure modes. The fuzzy AHP enabled systematic ranking of the variables, while the fuzzy TOPSIS algorithm provided easily understood and configurable computational procedures for ranking the failure modes based on optimal geometric paths to positive and negative ideal solutions.

**Conclusions:** The quality risk of ACTs could be reliably established using the fuzzy FMEA where aggregated experts’ decisions and risk variables’ weights are of considerable importance to the final ranking of quality failures.

## Introduction

Recent statistics published in the World Malaria Report have indicated that malaria remains endemic in Nigeria as it disproportionately bears an alarming 31.3% of the global burden of the disease [1]. Artemisinin-based combination therapy (ACTs) forms a principal component of World Health Organization (WHO) recommended malaria treatment regimens [2]. Safe and effective malaria prevention and eradication programmes strongly depend on the standards in the quality of ACTs [2]. The term quality is a broad term that summarizes multi-attribute criteria used to gauge the compliance level of the ACTs to fitness for intended clinical application throughout its lifecycle. ACTs critical product attributes include the drug’s identification, purity, potency, shelf-life, dissolution kinetics, bioequivalence, disintegration, and compliance with labelling and packaging requirements, in addition to several other pharmacopoeial and regional regulatory specification [3–6]. The quality of ACTs is essential to both patient safety during the use of the product and successful therapy.

Unfortunately, forensic analysis has revealed the structural vulnerability of the pharmaceutical supply chain (both formal and informal) of Low and Middle-income Countries (LMICs) to counterfeit and substandard ACTs posing significant public health risks [3,7,8]. The presence of low-quality ACTs greatly jeopardizes the standards of the supply chain resulting in diminished confidence in the healthcare systems [8]. Scientific evidence has shown that a compromised drug distribution system is a major setback with negative disruptive tendencies toward multifocal efforts in the fight against malaria in endemic regions. The proliferation of substandard and counterfeit ACTs is deemed economic sabotage that propels significant risks to parasite resistance, therapeutic failure, undesirable side effects, and even death [7,9].

An integrated approach to pharmaceutical product quality management is implemented through a standardised pharmaceutical quality system that is applicable throughout the product’s lifespan with a targeted goal of assuring patient safety and achieving therapeutic goals. Within the current regulatory thinking, as contextualized in the International Council on Harmonization guidelines (ICHQ9 and ICHQ10) quality risk assessment is indicated to ascertain compliance with the safety and efficacy standards of medicines throughout their lifecycle [10]. Accordingly, implementing the ICH Q9 principles in the antimalarial products supply chain assures continuous quality and process improvement plans with the ultimate goal of mitigating quality-dependent public health risks [11–13].

### Failure mode effect analysis

Failure mode effect analysis (FMEA) is a WHO-recommended quality risk assessment tool that is used in the quantitative or qualitative assessment of product, process, or system risks by linking the probability of occurrence (O) of fault (failure) with the severity (S) of the consequences of such fault, and its detectability (D) [14]. Since ICH Q9 guidelines have underscored the protection of patients by deploying a quality risk evaluation strategy [14], the derived quality risk information is crucial as it proffers an avenue for critical decision-making by regulators, industries, and all stakeholders in the ACT supply chain.

In the context of ACT quality risk assessment, the quality failure types that may potentially affect the ACT performance and ability to meet the desired target quality product profile to effectively treat malaria are assessed using the risk factors O, S, and D. In practice, for each failure mode, the scores of S, O, and D are rated by the cross-functional FMEA team members based on established FMEA risk factor scales [15,16]. A risk priority number (R.P.N) is then mathematically calculated as a simple product of the risk factors according to Equation 1, and the failure modes are prioritized in descending magnitude of the risk scores [4,13,17].

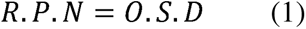

Thus, for an ACT product with *k* failure modes (FM), corrective actions and risk mitigation plans are proportionated based on hierarchy. Thus, FMEA troubleshoots non-conformances with critical quality requirements. Although this traditional FMEA (tFMEA) is simple to implement from a methodological context, it has been widely criticized for several cogent reasons [18,19]. Firstly, the risk factors’ weights are accorded equal importance, while in a practical sense, the inherent risk associated with each factor differs [16]. Consequently, equal R.P.N is often obtained for different values of the risk factors, confusing failure prioritization. Consider failure modes FM1, FM2, and FM3, each with (O, S, D) scores of (4, 7, 8), (4, 8, 7), and (8,7,4), respectively. Each of the failure modes would result in an RPN score of 224 resulting in non-discernible risks when Equation 1 is applied [18,19]. Thus, Equation 1 is itself mathematically controversial. Therefore, the reliability of the prioritized failure modes is also questionable. The implication of this is that failures that ought to have received the utmost corrective attention might become erroneously neglected or undetected, and the whole objective of the FMEA is defeated or compromised.

Another limitation of the tFMEA is that the use of crips values to represent various ratings of the risk factors does not adequately recapitulate human reasoning and natural decision-making culture. Human decisions are vague, and imprecise, albeit complete [20–22]. For instance, in rating the likelihood of occurrence of a failure mode, according to the crips value 5 to denote ‘moderate failure’ is not concordant with logical representation of human thinking and judgement [22,23]. Hence, crips values were considered insufficient in assignment of risk scores by FMEA decision team [18,19,22].

Considering the public health implications of healthcare decisions vis-à-vis their pharmacoeconomic impact, numerous modifications to the tFMEA approaches have been proposed. FMEA is considered a multi-attribute decision problem, hence Multicriteria Decision Analysis (MCDA) such as Analytical Hierarchical Process (AHP) and Technique for Order of Preference by Similarity to Ideal Solution (TOPSIS) have been independently and jointly integrated to increase the reliability and robustness of failure mode prioritization [18]. The concept of fuzzy logic was integrated into the MCDA tools to recapitulate FMEA team members’ intrapersonal and interpersonal divergence in logical reasoning [24].

Given the hitherto mentioned supply chain vulnerability, concerted efforts are vested by various governmental and non-governmental organizations as well as interested researchers to gauge the current status of ACTs. Medicines quality surveys are conducted as an external surveillance feedback mechanism to regulators, manufacturers, patients, and all stakeholders in the pharmaceutical supply chain [25]. The results of the survey may invoke a proactive risk mitigation plans systems for the management of product non-conformances, complaints, recalls, and continuous improvement plans [14]. The scientific literature on ACT quality assessment is fairly rich [3,26–28]. However, only a few studies adopted a risk-based approach in ACT quality risk analysis [4]. Moreover, the implementation of advanced FMEA analytics in quality assessment of ACTs remains elusive amidst the growing need for proactive pharmaceutical risk analysis in LIMCs, where both malaria and substandard antimalarials are endemic. This study therefore aims at providing a systematic methodology for implementing a hybrid FMEA based on fuzzy AHP integrated with fuzzy TOPSIS with a scope of strengthening the scientific quality of critical quality risk analysis of ACTs in counterfeit endemic regions.

The methodology was presented as follows. First, we briefly introduce the concept of fuzzy logic in the context of accounting for the expert’s vagueness in presenting their subjective opinions based on FMEA risk factor evaluation scales. In the latter part, we implemented a hybrid FMEA methodology based on fuzzy AHP and fuzzy TOPSIS in ACT failure mode prioritization problem by cross-functional FMEA team members. The paper also aims to communicate the computational utility of fuzzy logic, MCDA tools, and FMEA in a simplified and straightforward form. Therefore, derivations of formulae and details of mathematical theorems were out of the scope of the present work.

### Methodology for implementing hybrid fuzzy AHP and fuzzy TOPSIS in FMEA of ACTs

The urgent need for the fuzzy-based FMEA arose from an ongoing medicine quality survey that aims at critically evaluating the current status of ACTs marketed in the various strata of the pharmaceutical supply chains of three selected Northwestern states of Nigeria. The ethical approval for the conduct of the study was granted by the College Research Ethics Committee of the College of Health Sciences, Bayero University (Reference number: BUK/CHS-REC/121). Samples of different brands of ACTs were collected from each stratum and multiple quality attributes were analysed and documented using a modified Visual Tool for Quality Evaluation of Medicines. Pharmacopoeial tests were conducted by the quality control team according to USP41-NF 46. However, given the inherent limitations of the tFMEA highlighted above, a panel of quality experts was constituted to develop the hybrid fuzzy FMEA methodology. The historic quality control data of a blinded ACT sample was purposefully selected for this work

The cross-functional FMEA team comprises of five pharmaceutical quality experts with diverse expertise in pharmaceutical analysis, pharmaceutics and pharmaceutical technology, pharmaceutical engineering, pharmacology, and therapeutics. Following brainstorming the FMEA team members identified 15 failure modes. The team members recognized the limitations of the tFMEA and opted for a hybrid FMEA based on fuzzy AHP and fuzzy TOPSIS algorithms adopting the step-by-step procedure described in the subsequent sections.

### Formulation of the overall goal of the fuzzy FMEA model

From the analytical hierarchical structure (Fig 3), the overall goal of the FMEA was to prioritize the different failure modes of the ACT sample (Level 1). Level 2 represents the risk-defining variables (criteria), also called the risk factors (O, S, and D). Level 3 represents the various alternatives (failure modes in this case). In the fuzzy FMEA model, the scoring for the relative importance of the risk factors was independently provided by each FMEA team member by considering the crips scale/linguistic terms of judgement provided in Table 1. Similarly, the expert decision on each failure mode was independently provided by each FMEA team member using Table 2-4. To aid in the visualization and comprehension of the tables, the triangular membership functions (Fig 4-6) of the fuzzy numbers were created using Fuzzy Logic Designer in MATLAB (Version 9.13.0, R2022).

**Figure 1:**
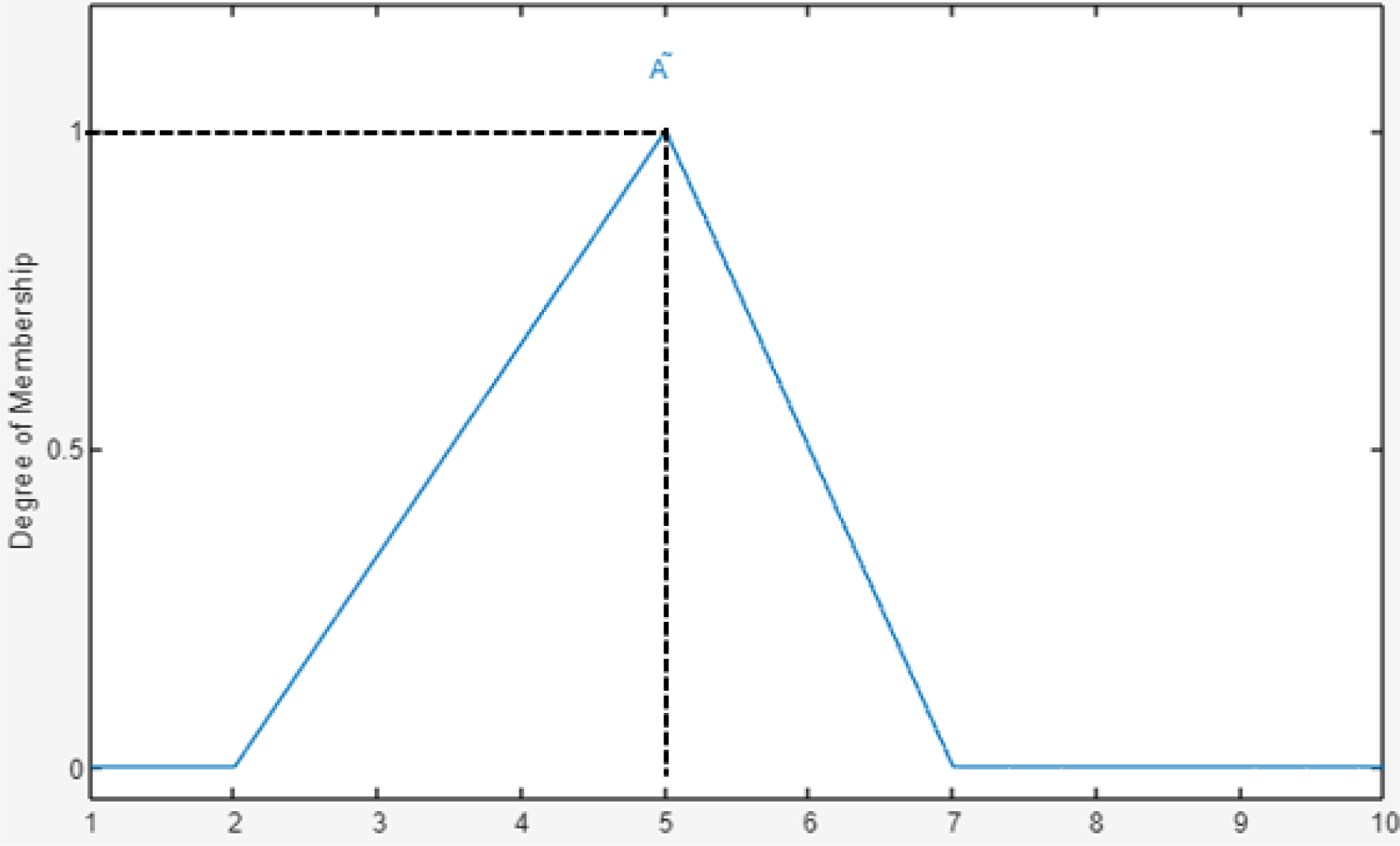
Triangular membership function. Here l=3, m=5, µ=7.

**Figure 2:**
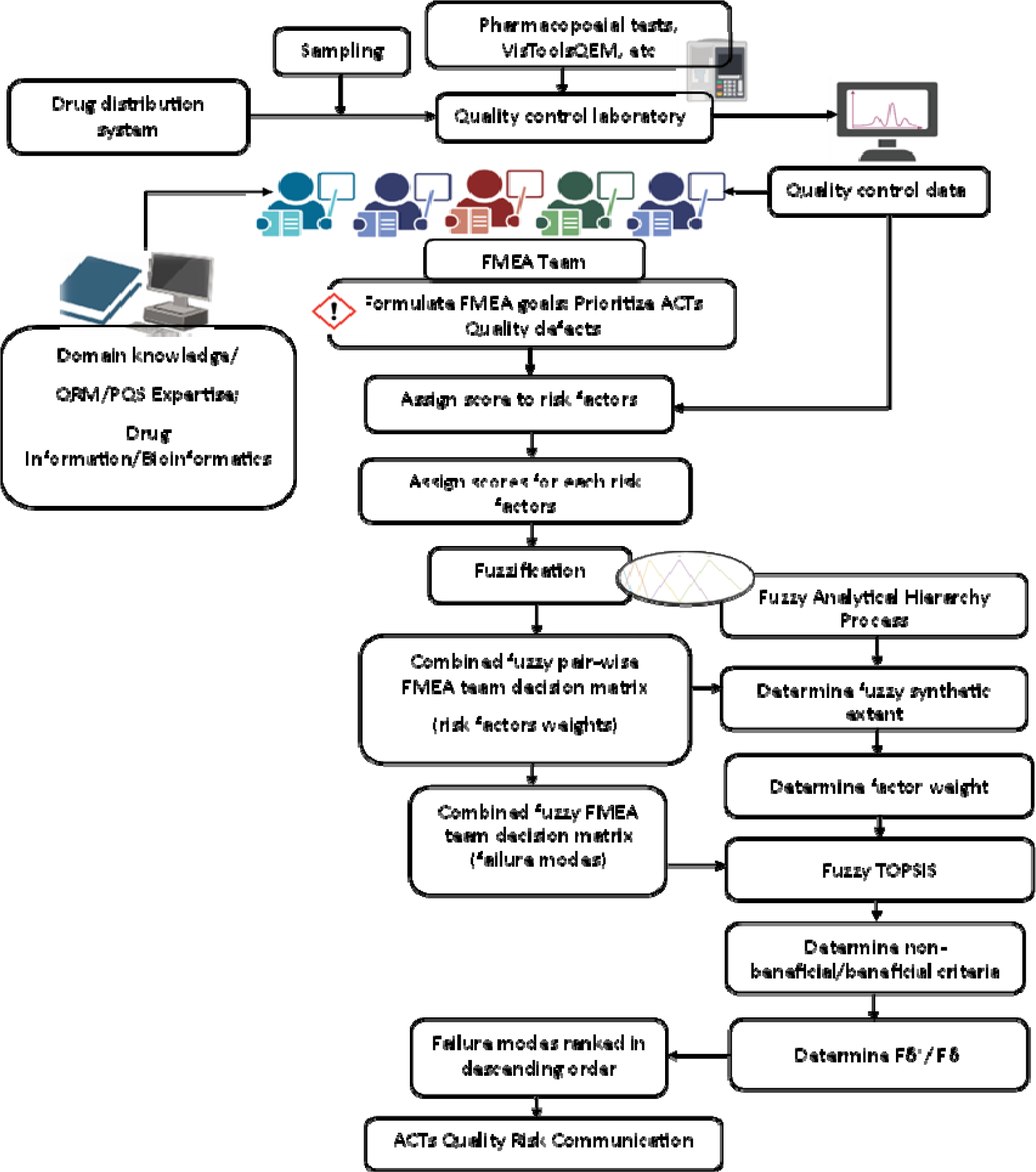
Systematic methodology for a hybrid fuzzy analytical hierarchy process and fuzzy TOPSIS in failure mode effect analysis of fixed-dose artemether-lumefantrine tablets. VisToolsQEM: Visual Tool for Quality Evaluation of Medicines, FMEA: Failure Mode Effect Analysis, PQS: Pharmaceutical Quality Systems, QRM: Quality Risk Management, TOPSIS: Technique for Order of Preference by Similarity to Ideal Solution. Fδ*: Fuzzy positive ideal solution, Fδ^-^: Fuzzy negative ideal solution.

**Figure 3:**
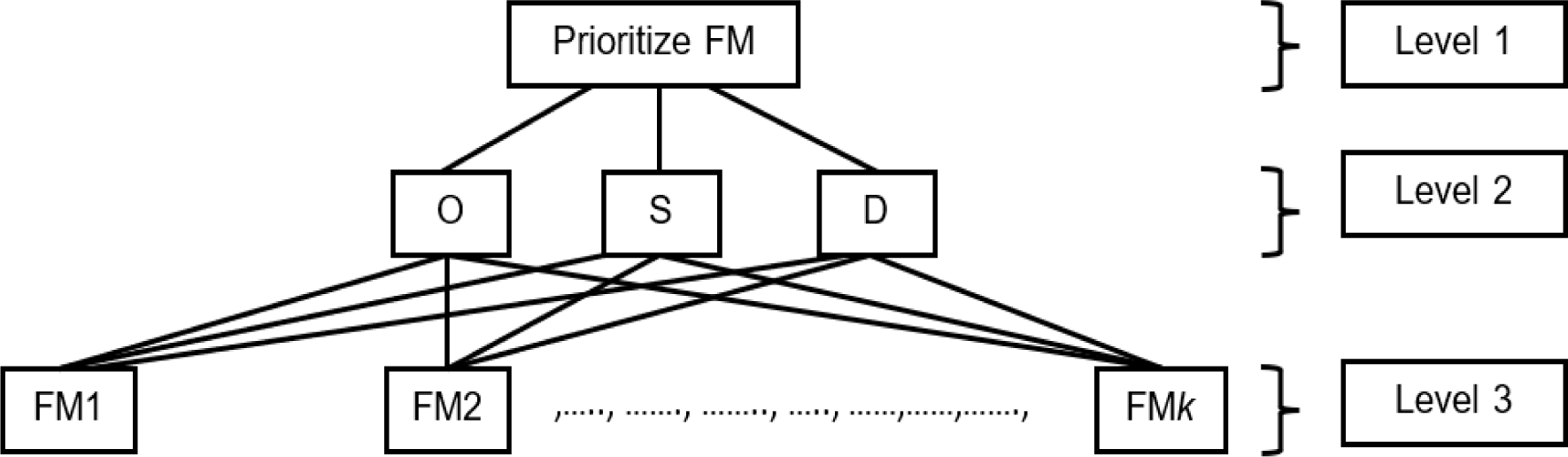
Analytical Hierarchy structure for failure mode prioritization. Level 1 is the goal of the Analytical Hierarchy Process, which is to prioritize the failure modes of the Artemether-Lumefantrine fixed-dose combination tablets. Level 2 represents the risk factors for probability of occurrence (O), Severity of failure (S), and likelihood of detection (D). Level 3 represents the various alternatives, which are all the fifteen failure modes.

**Figure 4:**
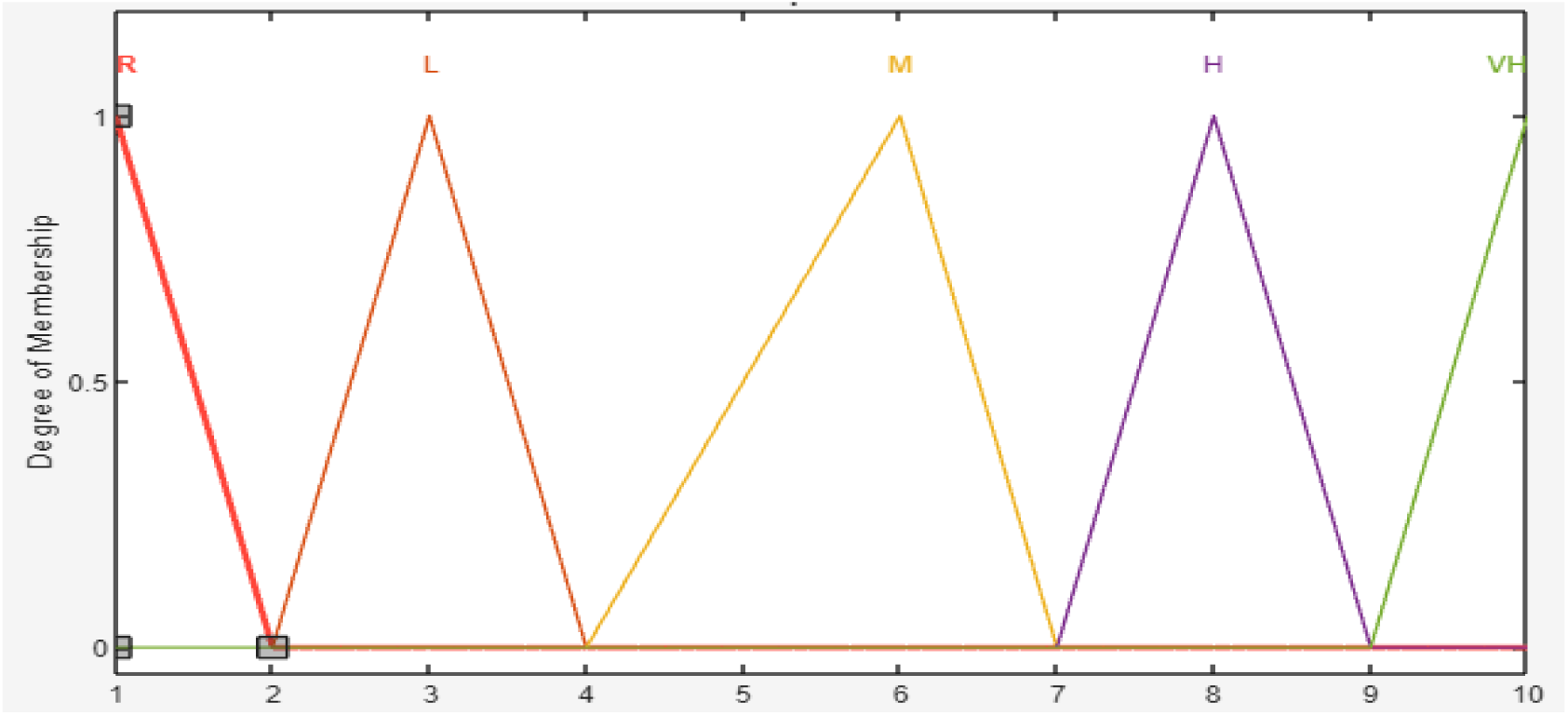
Membership function plot for Occurrence. R: Remote, L: Low, M: Moderate, H: High, VH; Very High.

**Figure 5:**
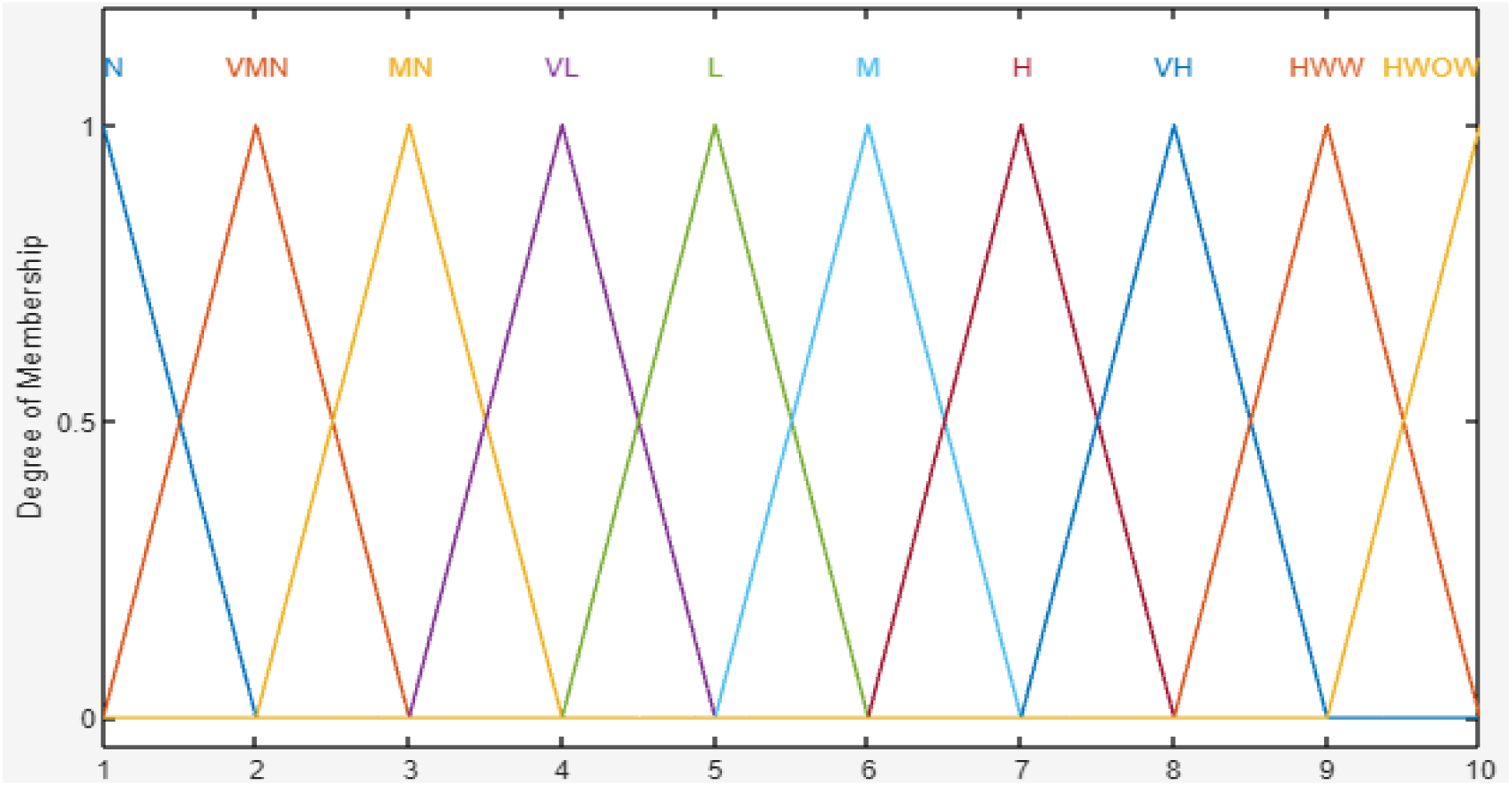
Membership functions for severity linguistic terms. N: None, VMN: Very minor, MN: Minor, VL: Very Low, L: Low, M: Moderate, H: High, VH; Very High, HWW: hazardous with warning: HWOW: Hazardous without warning.

**Figure 6:**
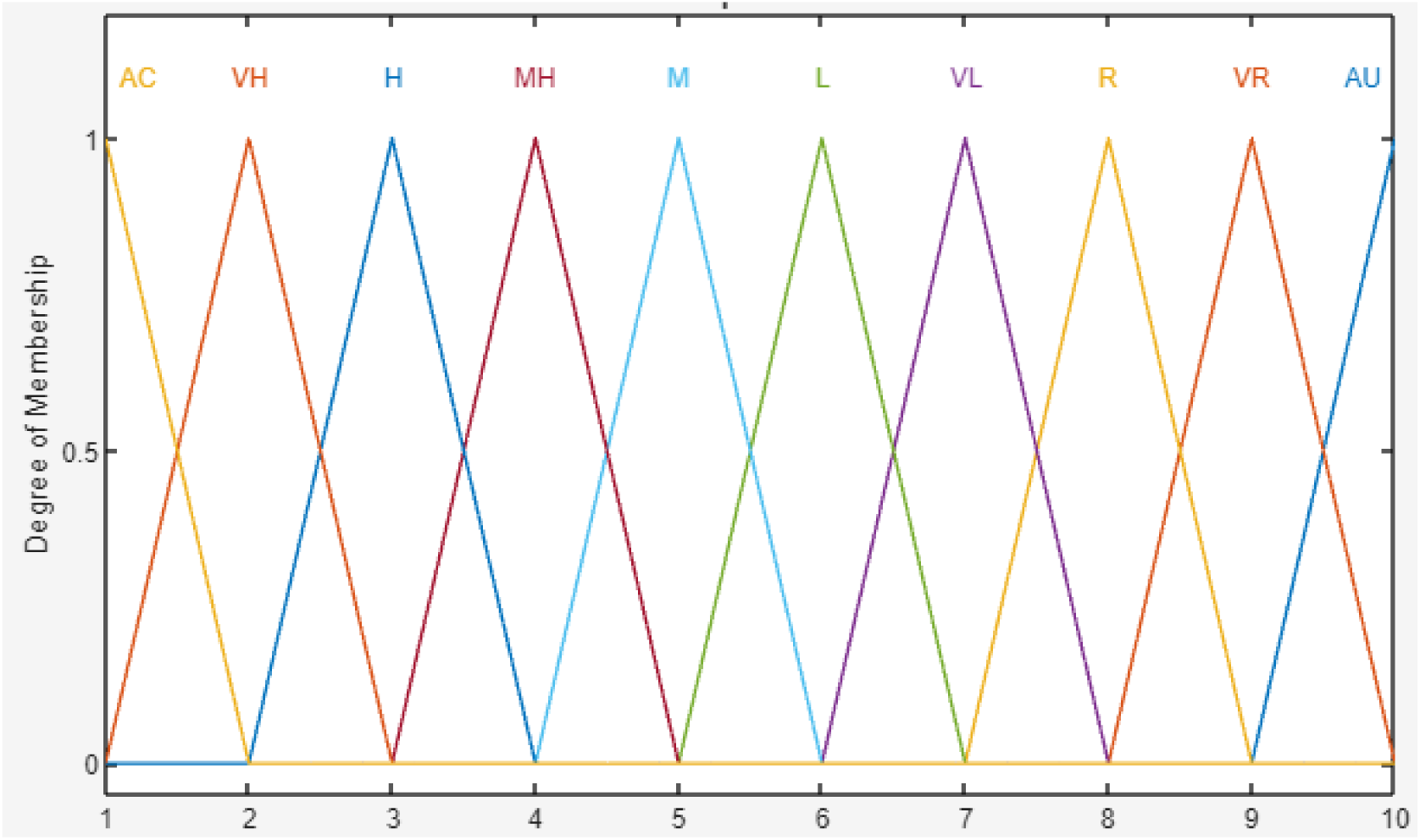
Membership functions for Detection linguistic terms. AC: Almost certain, Very Low, L: Low, M: Moderate, MH: Moderately high, H: High, VH; Very High, R: Remote, VR: Very remote, AU: Absolutely uncertain.

**Figure 7:**
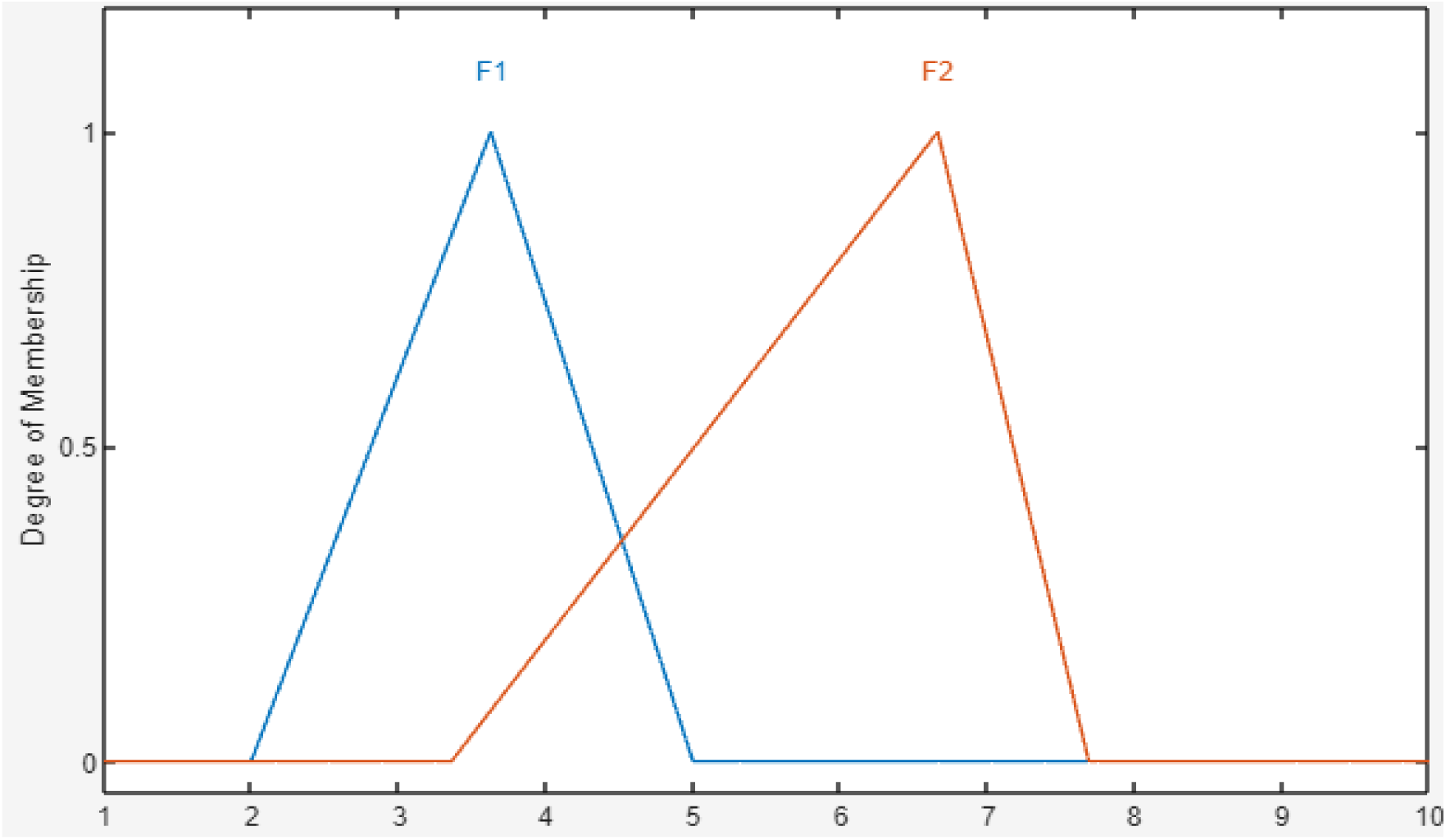
Comparison of fuzzy synthetic extents.

**Table 1:** Linguistic, crips, and fuzzy ratings for risk factor weighting.

| Linguistic term | Crips values | Fuzzy numbers |
| --- | --- | --- |
| Equal preference (EP) | 1 | (1, 1, 1) |
| Moderate preference (MP) | 3 | (2, 3, 4) |
| Strong preference (SP) | 5 | (4, 5, 6) |
| Very strong preference (VSP) | 7 | (6, 7, 8) |
| Extremely strong preference (EP) | 9 | (9, 9, 9) |

**Table 2:**
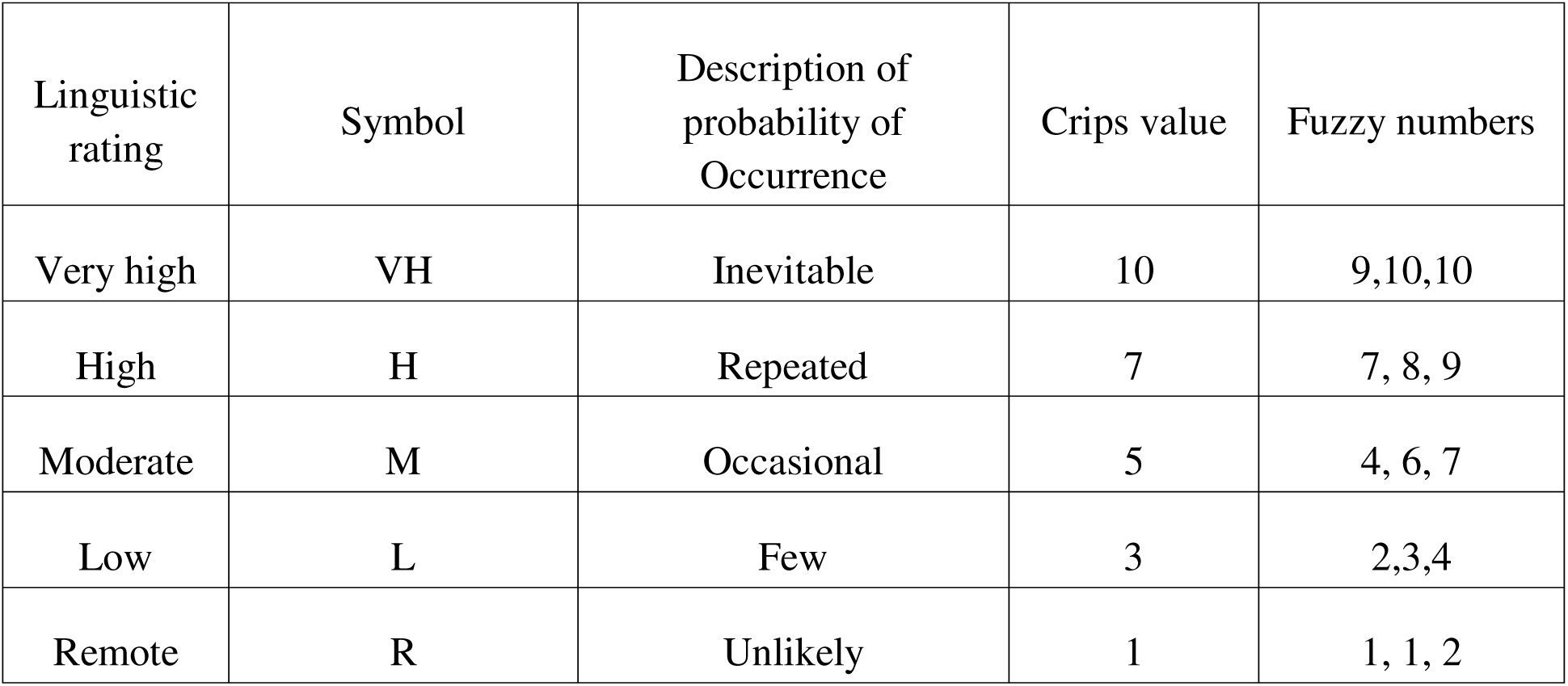
Linguistic, crips, and fuzzy ratings for occurrence [19].

| Linguistic rating | Symbol | Description of probability of Occurrence | Crips value | Fuzzy numbers |
| --- | --- | --- | --- | --- |
| Very high | VH | Inevitable | 10 | 9,10,10 |
| High | H | Repeated | 7 | 7, 8, 9 |
| Moderate | M | Occasional | 5 | 4, 6, 7 |
| Low | L | Few | 3 | 2,3,4 |
| Remote | R | Unlikely | 1 | 1, 1, 2 |

### Fuzzy analytical hierarchy process

In this method, we determined the weights of the risk factors using the extent analysis method [29,30]. The FMEA team members’ expert opinions of the three risk factors were provided using the linguistic ratings in Table 1 [18]. Each of the experts utilized his scientific judgment to value the risk factors. For convenience, we considered the crisp numeric values equivalent to the linguistic ratings. Also, to avoid ambiguity and inconsistency of decisions, a simple questionnaire was adopted from the literature (Supplementary file).

Let C be an *n*-ordered positive reciprocal matrix whose elements satisfy the condition [31]:

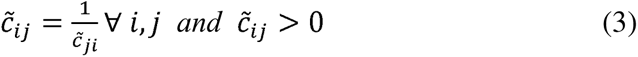

Then, the pairwise comparison matrix of the risk factors decision takes the form:

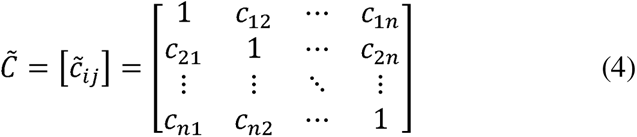

Also, Equation 5 describes a pairwise relation on the relative importance between two risk factors. The detail of the scale of relative importance was given in Table 1 [18].

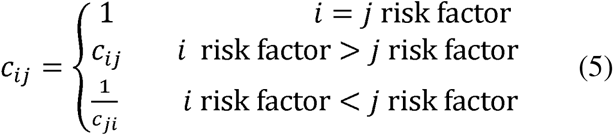

### Consistency analysis

To ensure that each expert opinion concerning risk factor weighting was consistent,

Saaty’s consistency ratio (CR) was utilized (Equation 6).

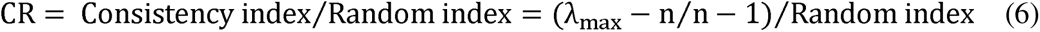

where λ_max_ is the principal eigenvalue of C. Because the matrix was a 3×3, a random considered for *CR*≤ 0.1 [31]. Inconsistent answers were sent back to the decision makers for index value of 0.58 calculated by Saaty was used. The acceptable consistent pairwise matrix was adjustment.

### Fuzzification

After obtaining a consistent pairwise comparison matrix, the crips numeric decision of the experts were converted into the corresponding fuzzy numbers.

Let *A* be a crips numeric value, and *Ã* be the corresponding fuzzy set whose elements are (*l, m, μ*) which represents the vagueness of the expert decision. The optimum certainty of the expert is represented by *m*, while the associated lower and upper deviations were respectively denoted by *l* and μ,[18]. The aggregated fuzzified pairwise comparison matrix was computed as:

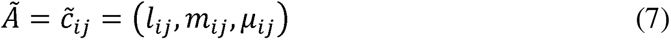

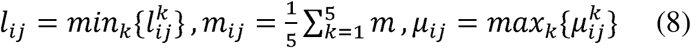

The general arithmetic operations with two positive triangular fuzzy numbers, F_1_ and F_2_, are shown in Table 5 [18,19]. These general operations were used throughout the calculations.

**Table 3:**
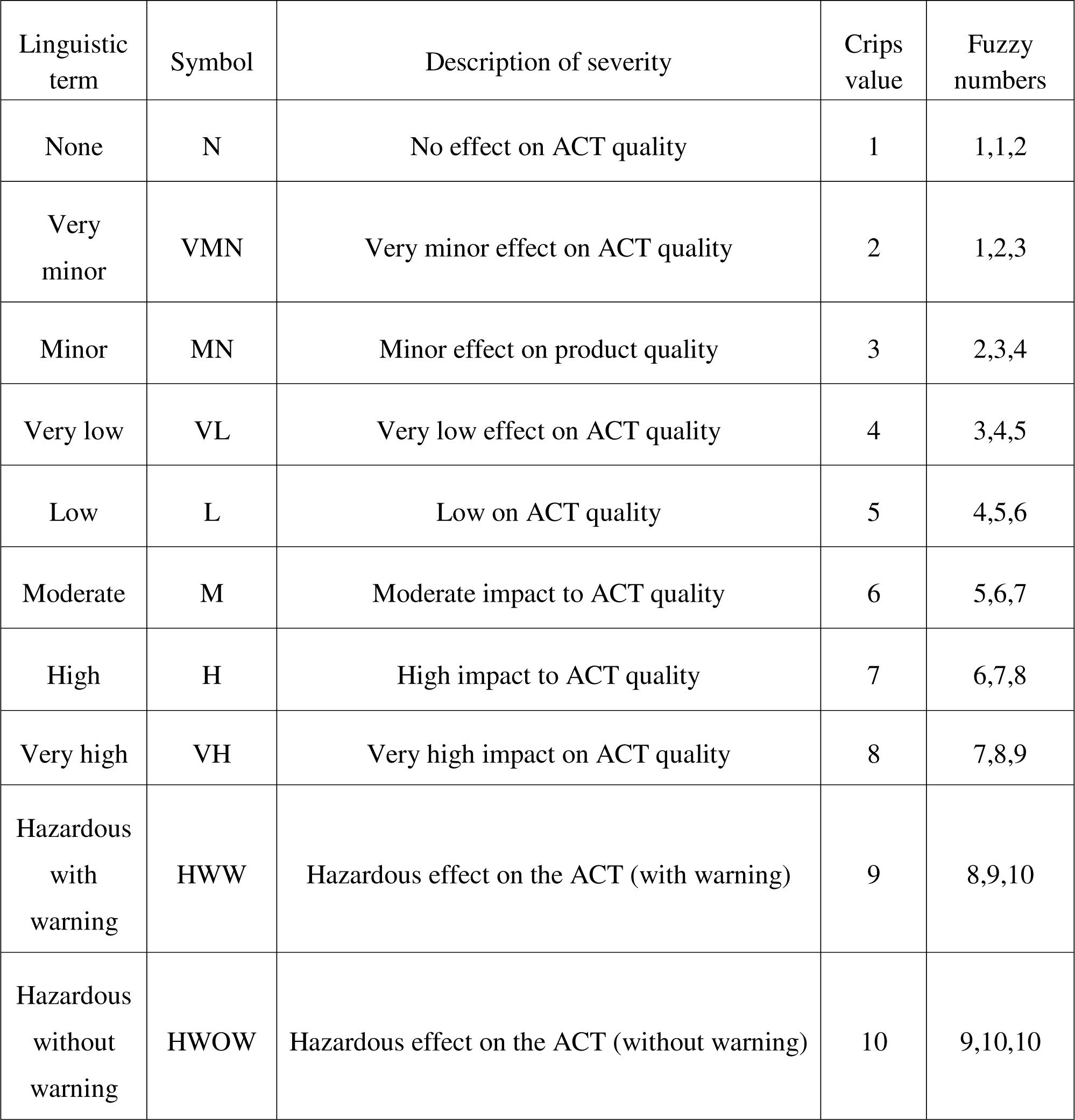
Linguistic, crips, and Fuzzy ratings for Severity.[19].

| Linguistic term | Symbol | Description of severity | Crips value | Fuzzy numbers |
| --- | --- | --- | --- | --- |
| None | N | No effect on ACT quality | 1 | 1,1,2 |
| Very minor | VMN | Very minor effect on ACT quality | 2 | 1,2,3 |
| Minor | MN | Minor effect on product quality | 3 | 2,3,4 |
| Very low | VL | Very low effect on ACT quality | 4 | 3,4,5 |
| Low | L | Low on ACT quality | 5 | 4,5,6 |
| Moderate | M | Moderate impact to ACT quality | 6 | 5,6,7 |
| High | H | High impact to ACT quality | 7 | 6,7,8 |
| Very high | VH | Very high impact on ACT quality | 8 | 7,8,9 |
| Hazardous with warning | HWW | Hazardous effect on the ACT (with warning) | 9 | 8,9,10 |
| Hazardous without warning | HWOW | Hazardous effect on the ACT (without warning) | 10 | 9,10,10 |

**Table 4:** Linguistic, crips, and Fuzzy ratings for Detection.[19].

| Linguistic rating | Symbol | Description of likelihood of detection | Crips value | Fuzzy numbers |
| --- | --- | --- | --- | --- |
| Absolute uncertainty | AU | Design control will not and/or cannot detect potential failure. | 10 | 9, 10, 10 |
| Very remote | VR | The chance that the design control will detect potential failure is very remote | 9 | 8, 9, 10 |
| Remote | R | The chance that the design control will detect potential failure is remote | 8 | 7, 8, 9 |
| Very low | VL | The chance that the design control will detect potential failure is very low | 7 | 6, 7, 8 |
| Low | L | The chance that the design control will detect potential failure is low | 6 | 5, 6, 7 |
| Moderate | M | The chance that the design control will detect potential failure is moderate | 5 | 4, 5, 6 |
| Moderately high | NH | The chance that the design control will detect potential failure I moderately high | 4 | 3, 4, 5 |
| High | H | The chance that the design control will detect potential failure is high | 3 | 2, 3, 4 |
| Very high | VH | The chance that the design control will detect potential failure is very high | 2 | 1, 2, 3 |
| Certain | C | The chance that the design control will detect potential failure is certain | 1 | 1, 1, 2 |

**Table 5:** General arithmetic operations for calculations with positive triangular fuzzy numbers.

| Operation | Applied fuzzy operation |
| --- | --- |
| Addition | $\tilde{F}_1 \oplus \tilde{F}_2 = (l_1, m_1, \mu_1) \oplus (l_2, m_2, \mu_2) = (l_1 + l_2, m_1 + m_2, \mu_1 + \mu_2)$ |
| Multiplication | $\tilde{F}_1 \otimes \tilde{F}_2 = (l_1, m_1, \mu_1) \otimes (l_2, m_2, \mu_2) = (l_1 \times l_2, m_1 \times m_2, \mu_1 \times \mu_2)$ |
| Division | $\tilde{F}_1 \div \tilde{F}_2 \approx (\frac{l_1}{\mu_2}, \frac{m_1}{m_2}, \frac{\mu_1}{l_2})$ |
| Subtraction | $\tilde{F}_1 - \tilde{F}_2 = (l_1 - l_2, m_1 - m_2, \mu_1 - \mu_2)$ |

### Computation of fuzzy synthetic extent

The fuzzy synthetic extent (*F_i_*) with respect to each risk variable was then calculated as in Equation 9 [18,29].

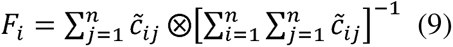

Secondly, the degree of superiority of the fuzzy synthetic extents was calculated according to Equation 10 [29,30]. Given that there were three risk factors, six degrees of superiorities were compared: *V*(F_1_≥F_2_), *V*(F_1_≥F_3_), *V*(F_2_≥F_1_), *V*(F_2_≥F_3_), *V*(F_3_≥F_1_), *V*(F_3_≥F_2_)

Generally, the degree of possibility that F_1_≥F_2_was given by Equation 10 [18].

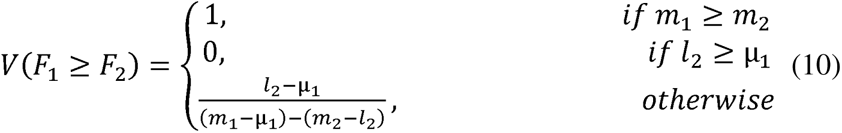

Hence, the degree of possibility of a given convex fuzzy number to be superior than *k* convex fuzzy numbers was calculated using Equation 11. Accordingly, the weight vector and it’s normalized for were determined according to Equation 12 and 13, respectively [29,30].

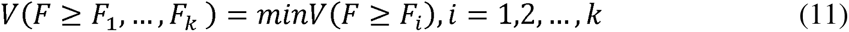

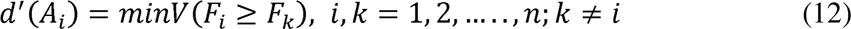

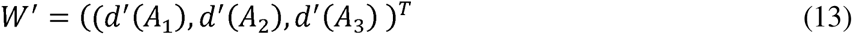

### Fuzzy TOPSIS expert assessment of failure modes

On the basis of the historic quality analysis data of the ACT sample, the cross-functional team members provided subjective assessment of each failure mode. Compliance of pharmacopoeial specifications were considered as benchmark for assessment of optimum compliance to quality standards. The risk posed to the ACT quality was subjectively evaluated based on deviation from pharmacopoeial tolerance limits of the investigated quality attributes.

This was supplemented by regional regulatory requirements to critical quality requirements where applicable. Again, each team member was instructed to use linguistic terms, which in our case were equivalent to the crisps numeric values for ranking the occurrence, severity, and detection based on Table 2, 3, and 4, respectively.

The crisp ratings were converted to corresponding fuzzy numbers and the aggregated subjective group opinion matrix computed using Equation 14 [22].

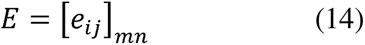

### Normalization of the combined/aggregated fuzzy decision matrix

Sets of beneficial (ξ) and cost (ς) risk factors were assigned based on the synthesized risk variable weights. We hypothetically considered the low-weighted risk factors as the ξ,, while the high-weighted risk factor as the ς. Normalization of the aggregated FMEA opinion proceed according to Equation 15 and 17, respectively [18,32–35].

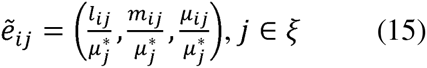

where,

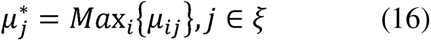

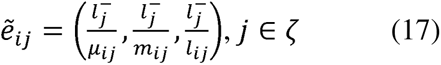

where,

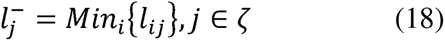

### Computation of weighted normalized fuzzy decision matrix

To obtain the normalized fuzzy decision matrix, the weight vectors were multiplied with fuzzy decision matrix.

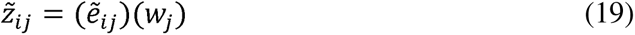

### Determination of Fuzzy Positive Ideal Solution (F*) and Fuzzy Negative Ideal Solution (F^-^)

The fuzzy positive ideal solution and fuzzy negative ideal solution were identified using Equation 20 and 21, respectively [32,35].

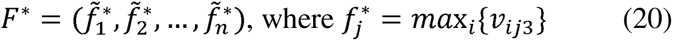

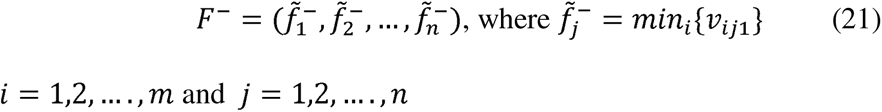

In general terms, the distance between two triangular fuzzy numbers α and β,can be calculated using Equation 22 [35]. Accordingly, the distances of each failure mode from fuzzy positive ideal solution and fuzzy negative ideal solution were calculated using Equation 23 and 24, respectively [18,33,35].

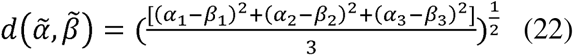

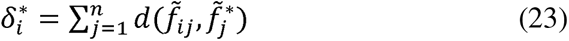

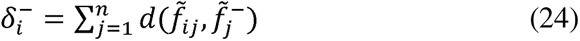

### Closeness coefficient

On the basis of Equation 24, failure mode a failure mode was deemed closer to *F*\*, and distant from *F*^-^ as the closeness coefficient tends to unity [35].

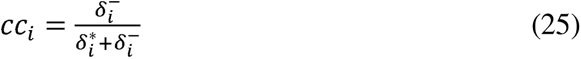

## Results and discussion

### Fuzzy AHP for Group analysis of the relative importance of the risk variables

In FMEA critical evaluation of quality risk of pharmaceutical products is a collective responsibility of the cross-functional quality experts whose opinions on the comparative weights of the risk-determining variables were highly subjective. The analytical hierarchy process (AHP) is a powerful technique for establishing the degree of comparative superiority between criteria in practical MCDA [29,30]. The risk defining variables in the tFMEA are not equally important, and treating them as such in risk priority calculation would result in non-reliable results. With the help of AHP the hierarchical connections of the risk variables can be analysed and their relative weights defined. However, AHP does not factor in the vagueness of the cross functional team members in making pairwise comparison. By integrating fuzzy logic concept into AHP, this problem could be adequately resolved [18,24,29].

In this study, the team members utilized their professional and scientific judgements to linguistically rank the three risk variables in a pairwise form. To resolve the uncertainties or impreciseness associated with subjective expert assessment of the risk factors, each linguistic term (denoted by the crisp numeric value) was converted in to the corresponding triangular fuzzy number. In this form, the subjectivities of the experts concerning their perception of the risk factors is numerically presented and conceptually appreciated. At this point, the uncertainty and impreciseness were apparent as all the pairwise comparison matrices were non-identical (Table 6). This was more evident in the individual and aggregated fuzzified pairwise comparison matrix (Table 7-8). However, throughout each expert decision matrix, transitivity rule was observed to ensure that pairwise relations of the risk variables were not contradictory. Inconsistent pairwise comparison results in errors in the computation of the aggregated hierarchical weights with consequences on the reliability of the final failure mode prioritization. Therefore, to ensure consistent judgement, each pairwise comparison matrix was analysed using Saaty’s CR [31]. Initially, there were two situations with inconsistent judgement (CR>>0.1), but the questionnaire was sent back to the concerned TM to rectify the situation. Finally, in all expert opinions, a value of CR<0.1 was obtained before proceeding to aggregate the individual fuzzified matrices into fuzzy group decision matrix. The aggregated fuzzified expert opinion formed the group opinion of the risk factor weights for subsequent extent analysis.

**Table 6:**
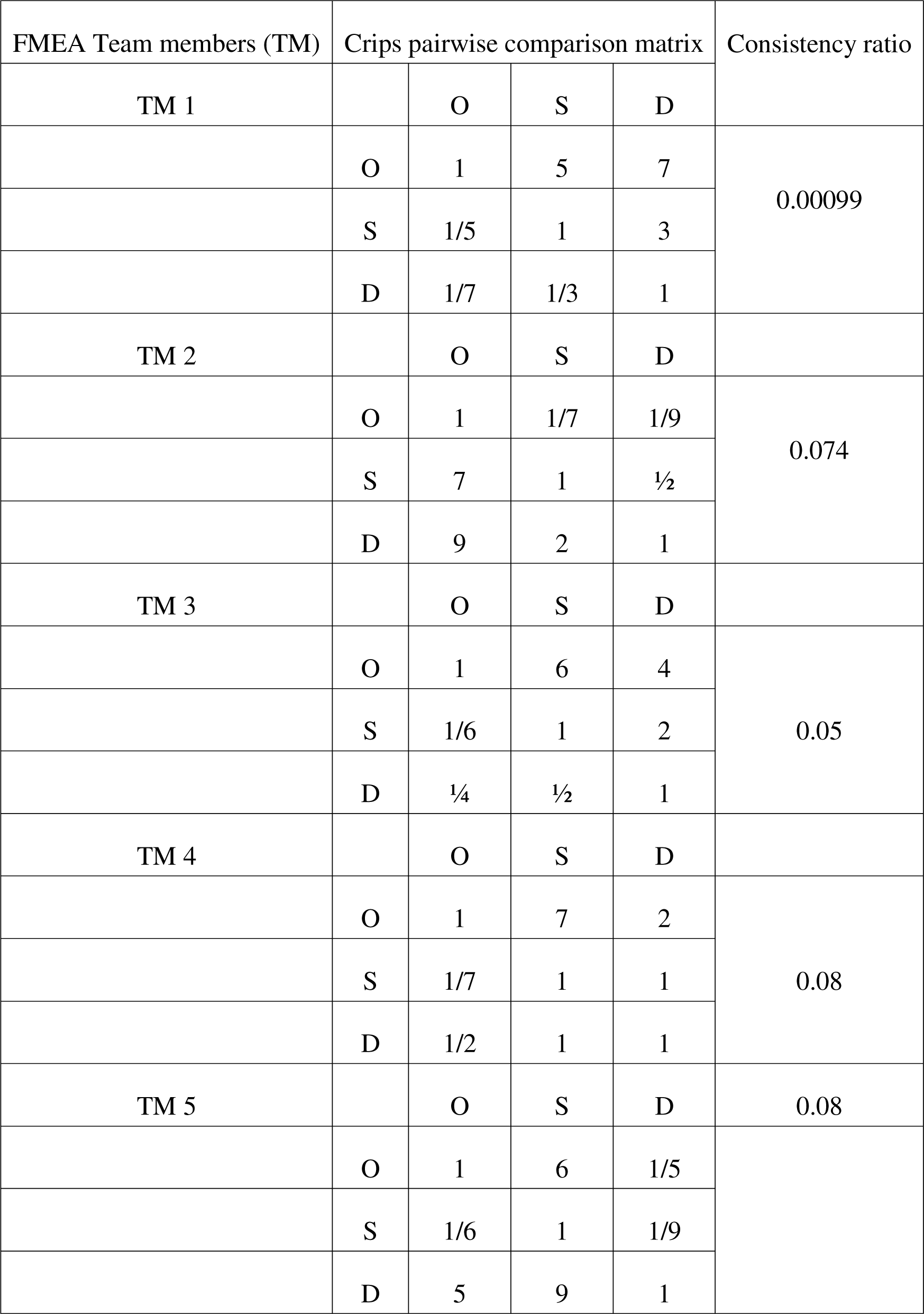
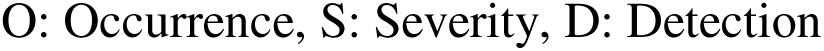
Pairwise comparison matrix of risk factors (crisps values) of each cross functional team member.

| FMEA Team members (TM) | Crips pairwise comparison matrix |  |  |  | Consistency ratio |
| --- | --- | --- | --- | --- | --- |
| TM 1 |  | O | S | D |  |
|  | O | 1 | 5 | 7 | 0.00099 |
|  | S | 1/5 | 1 | 3 |  |
|  | D | 1/7 | 1/3 | 1 |  |
| TM 2 |  | O | S | D | 0.074 |
|  | O | 1 | 1/7 | 1/9 |  |
|  | S | 7 | 1 | 1/2 |  |
|  | D | 9 | 2 | 1 |  |
| TM 3 |  | O | S | D | 0.05 |
|  | O | 1 | 6 | 4 |  |
|  | S | 1/6 | 1 | 2 |  |
|  | D | 1/4 | 1/2 | 1 |  |
| TM 4 |  | O | S | D | 0.08 |
|  | O | 1 | 7 | 2 |  |
|  | S | 1/7 | 1 | 1 |  |
|  | D | 1/2 | 1 | 1 |  |
| TM 5 |  | O | S | D | 0.08 |
|  | O | 1 | 6 | 1/5 |  |
|  | S | 1/6 | 1 | 1/9 |  |
|  | D | 5 | 9 | 1 |  |

O: Occurrence, S: Severity, D: Detection

**Table 7:**
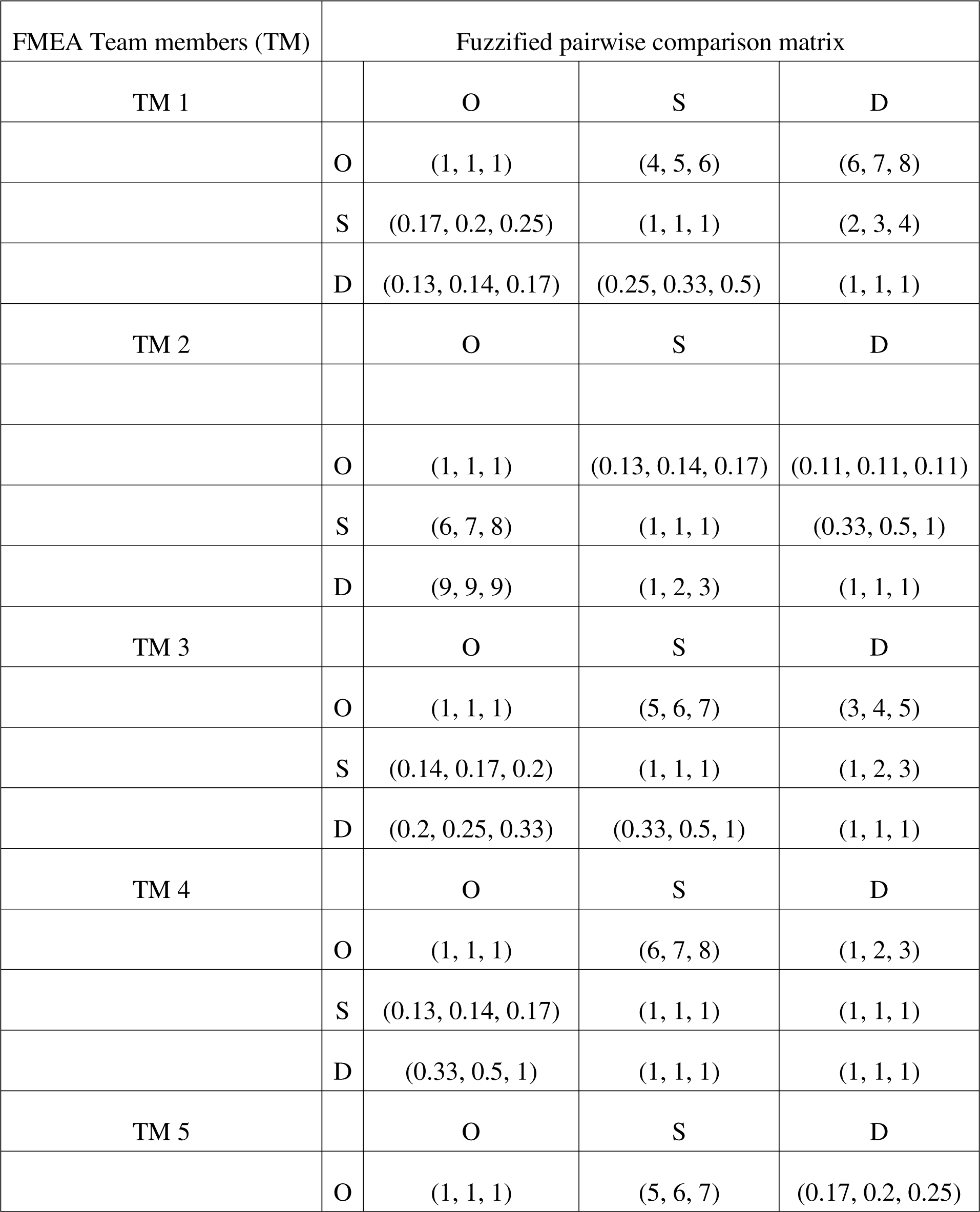
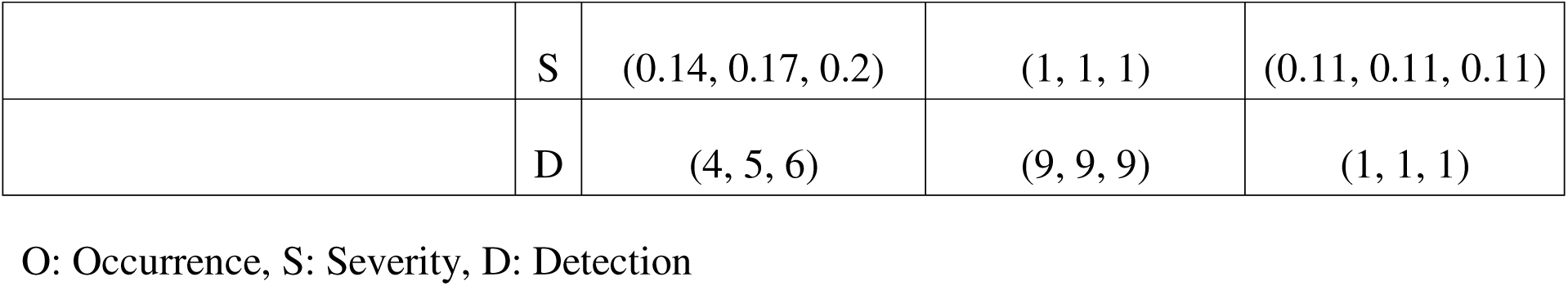
Fuzzified pairwise comparison matrix of risk factors by individual cross functional team members.

| FMEA Team members (TM) | Fuzzified pairwise comparison matrix |  |  |  |
| --- | --- | --- | --- | --- |
| TM 1 |  | O | S | D |
|  | O | (1, 1, 1) | (4, 5, 6) | (6, 7, 8) |
|  | S | (0.17, 0.2, 0.25) | (1, 1, 1) | (2, 3, 4) |
|  | D | (0.13, 0.14, 0.17) | (0.25, 0.33, 0.5) | (1, 1, 1) |
| TM 2 |  | O | S | D |
|  | O | (1, 1, 1) | (0.13, 0.14, 0.17) | (0.11, 0.11, 0.11) |
|  | S | (6, 7, 8) | (1, 1, 1) | (0.33, 0.5, 1) |
|  | D | (9, 9, 9) | (1, 2, 3) | (1, 1, 1) |
| TM 3 |  | O | S | D |
|  | O | (1, 1, 1) | (5, 6, 7) | (3, 4, 5) |
|  | S | (0.14, 0.17, 0.2) | (1, 1, 1) | (1, 2, 3) |
|  | D | (0.2, 0.25, 0.33) | (0.33, 0.5, 1) | (1, 1, 1) |
| TM 4 |  | O | S | D |
|  | O | (1, 1, 1) | (6, 7, 8) | (1, 2, 3) |
|  | S | (0.13, 0.14, 0.17) | (1, 1, 1) | (1, 1, 1) |
|  | D | (0.33, 0.5, 1) | (1, 1, 1) | (1, 1, 1) |
| TM 5 |  | O | S | D |
|  | O | (1, 1, 1) | (5, 6, 7) | (0.17, 0.2, 0.25) |
|  | S | (0.14, 0.17, 0.2) | (1, 1, 1) | (0.11, 0.11, 0.11) |
|  | D | (4, 5, 6) | (9, 9, 9) | (1, 1, 1) |

O: Occurrence, S: Severity, D: Detection

**Table 8:** Aggregated Fuzzified pair-wise comparison matrix of all cross functional team members.

| Risk factors | O | S | D | Normalized weight vector |
| --- | --- | --- | --- | --- |
| O | (1, 1, 1) | (0.125, 4.83, 8) | (0.11, 2.66, 8) | 0.43 |
| S | (0.13, 1.54, 8) | (1, 1, 1) | (0.11, 1.32, 4) | 0.28 |
| D | (0.13, 2.97, 9) | (0.25, 2.57, 9) | (1, 1, 1) | 0.29 |

In the latter part of the fuzzy AHP extent analysis, a pairwise analysis of the degree to which risk factor *i* outweighs risk factor j was computed. Thus, based on Equation 9 the computed fuzzy synthetic were as follows:

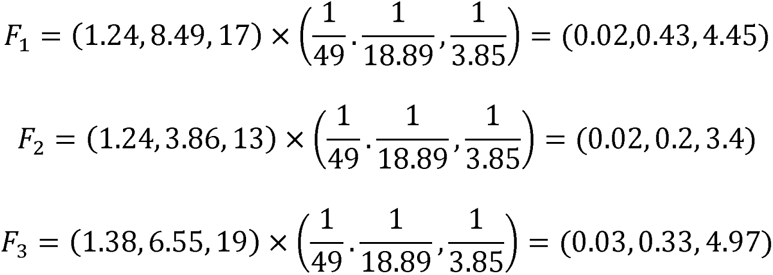

Similarly, Equation 10 provided a conditional relation for determining the degrees of possibilities for fuzzy synthetic extent i to be greater than j as: V (F1 ≥ F2) = 1, V(F1 ≥ F3) = 1, V(F2 ≥ F1) = 0.6452, V(F2 ≥ F3) = 0.7463, V(F3 ≥ F1) = 0.6582, V(F3 ≥ F2) =1.

Hence, after comparisons of the degree of possibilities it was found that V(F1 ≥ F2, F3)1, V(F2 ≥ F1,F3) = 0.6452, V(F3 ≥ F1,F2) = 0.6582. Finally, the normalized weight vectors were *W*^0^ = 0.43, *W^S^* = 0.28, and *W*^D^ = 0.29 for O, S, and D, respectively.

The normalized risk variable weights indicated that the risk variable O was more important than S and D. On this basis, O was hypothetically assigned as non-beneficial criteria and their minimum value was desired in the risk computation. Although S was slightly greater than D, both risk factors were designated as beneficial criteria, and hence their maximum values were desired.

### Fuzzy TOPSIS ranking of the failure modes

There were 15 failure modes each of which received an O, S, and D risk scores by each cross-functional team member based on the scales provided in Table 2, 3, and 3, respectively. The crips numeric value for each risk score indicates the team member’s expert judgement on that factor. It was also evident that, the inherent intrapersonal subjectivities were not captured by the crips values, in contrast to the fuzzified risk score based on the triangular fuzzy numbers. The aggregated group decision of all risk defining variable for all failure modes was presented in Table 8.

Unlike the fuzzy AHP analysis, the TOPSIS had the advantage of circumventing the need for pairwise comparison, which would otherwise make the individual and aggregated decisions matrices cumbersome. Also, there were no priory requirement for assuring transitivity rule and consistency ratios as well as the need for computation of fairly large number of fuzzy synthetic extents and their degree of relations.

The greatest advantage of TOPSIS utilized in this study was in rankings the large numbers of failure modes. Accordingly, the fuzzy TOPSIS was geared towards capturing the vagueness of the team members opinions. To achieve this, we explored the fundamental principles of TOPSIS where alternatives were ranked on the based on the Euclidean distances from hypothetical ideal and non-ideal solutions.

In FMEA, the higher the risk factor score, the more the risk priority number [19]. Hence, in our study we conditioned the risk variable with highest group weight of importance as the non-beneficial criteria. Conversely, the low weight bearing factors were regarded as beneficial criteria. Following determination of the closeness coefficient, the ranking of the failure modes in descending order of risk priorities were FM9≻ FM12≻ FM13≻ FM10≻ FM11≻ FM2≻ FM4≻ FM6≻ FM3≻ FM1≻ FM5≻ FM7≻FM8≻ FM15≻ FM14 (Table 10).

**Table 9:** Aggregated fuzzified ratings of the failure modes.

| Failure mode code | Occurrence | Severity | Detection |
| --- | --- | --- | --- |
| FM1 | (1, 1.8, 4) | (1, 2.8, 9) | (1, 3.2, 10) |
| FM2 | (1, 4.5, 10) | (1, 4.5, 10) | (1, 4.25, 9) |
| FM3 | (1, 3, 7) | (1, 3.6, 10) | (1, 2.8, 7) |
| FM4 | (2, 5.4, 9) | (2, 5.4, 10) | (1, 4.4, 7) |
| FM5 | (2, 5.2, 7) | (1, 5, 9) | (1, 3, 6) |
| FM6 | (1, 3.6, 7) | (1, 4, 8) | (1, 3.6, 10) |
| FM7 | (1, 4.6, 10) | (1, 4.2, 10) | (1, 2, 5) |
| FM8 | (1, 4.2, 10) | (1, 4.4, 10) | (1, 1.8, 5) |
| FM9 | (4, 8.2, 10) | (7, 9.2, 10) | (1, 4.6, 10) |
| FM10 | (1, 3.2, 10) | (1, 3, 10) | (1, 4.8, 10) |
| FM11 | (1, 3.2, 10) | (1, 3, 10) | (1, 4.6, 10) |
| FM12 | (4, 8.6, 10) | (6, 8.8, 10) | (1, 5.2, 10) |
| FM13 | (2, 6.2, 10) | (2, 6.6, 10) | (1, 5.4, 10) |
| FM14 | (1, 3.6, 10) | (1, 4.2, 10) | (1, 1.6, 4) |
| FM15 | (1, 3.8, 10) | (1, 4, 10) | (1, 1.6, 3) |

**Table 10:**
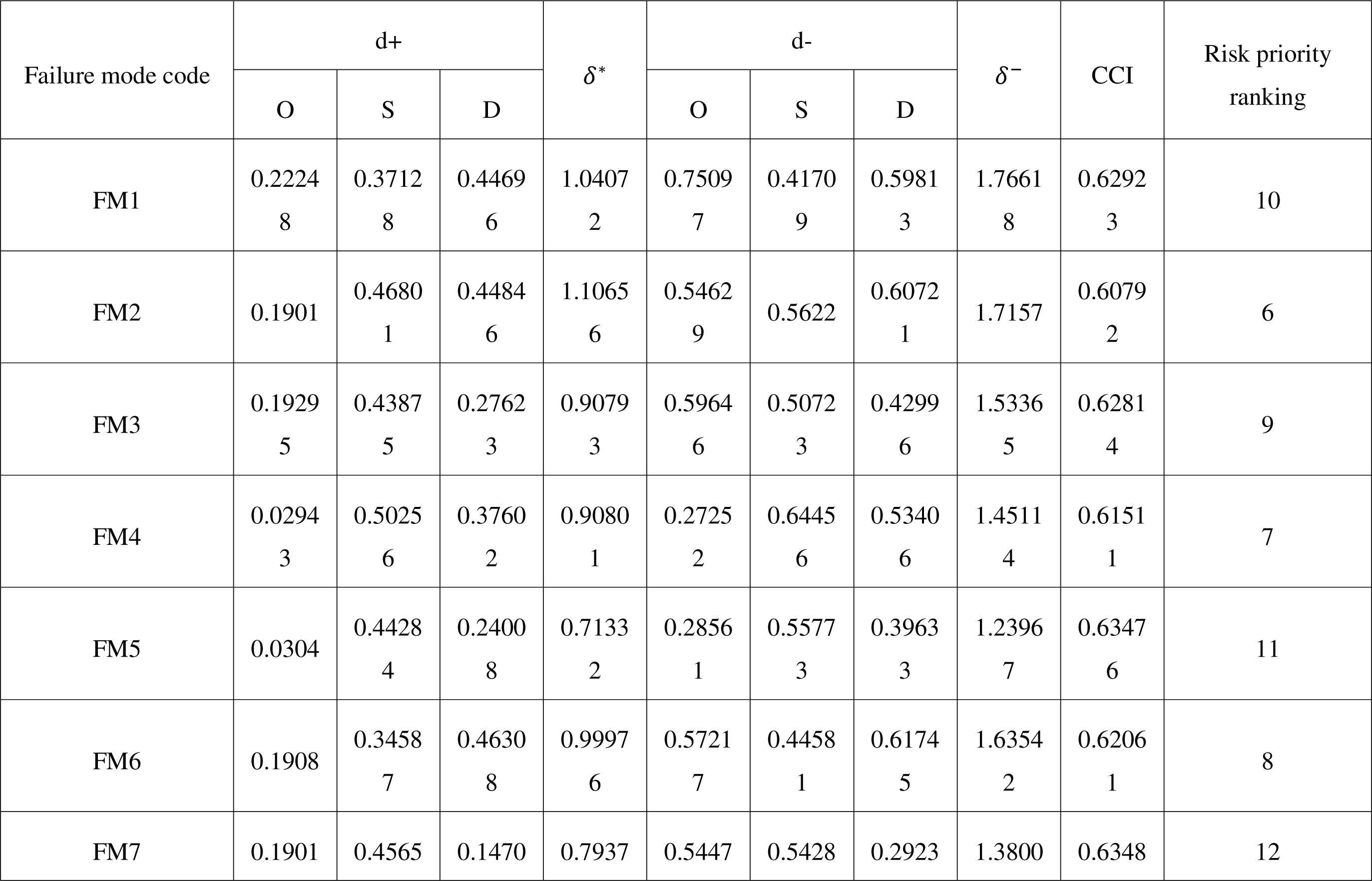
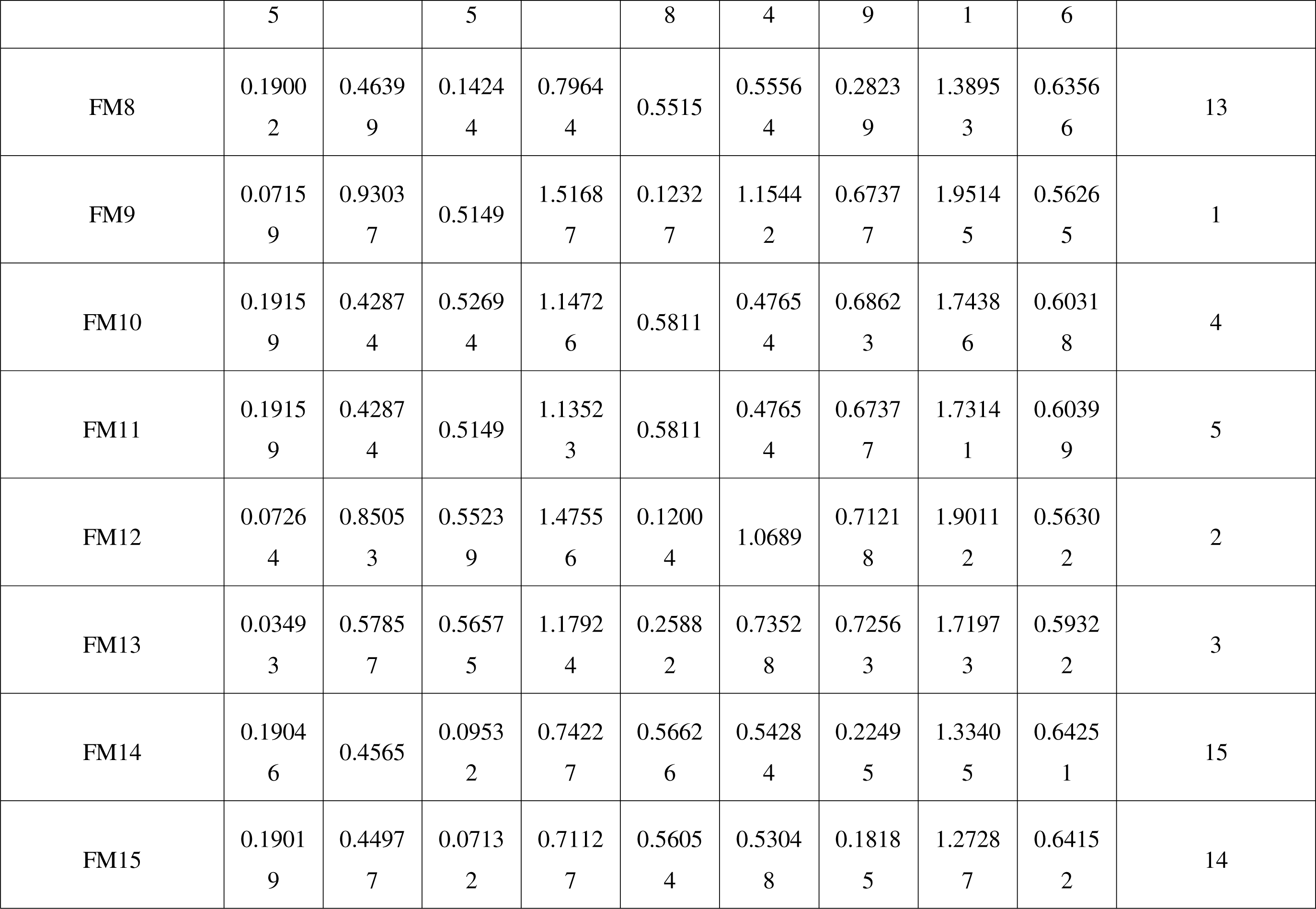
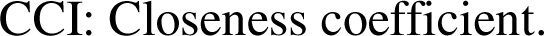
Closeness coefficient and final failure modes ranking.

**Table 11:** Description of failure modes and the final ranking.

| Failure mode | Failure mode description | Risk priority ranking |
| --- | --- | --- |
| FM1 | Failed mobile authentication service (MAS). There was no feedback when the code was sent for authentication via short message service. | 10 |
| FM2 | Inappropriate primary packaging/Containers not properly sealed/. | 6 |
| FM3 | Labelling issues. Legibility and indelibility of label information; Trade name misspelled/Inconsistent spelling of API/Conformity to registered drug. Leaflet explaining dosage, the drug content, the adverse effects, the drug actions, and how the drug should be taken absent in the product (Also not printed on the secondary pack). Smudge proof. Legibility and indelibility of the information. | 9 |
| FM4 | Hologram/Logo not authentic (e.g does not change when viewed from different angle | 7 |
| FM5 | Tablets not coated (Sugar coating, enteric coating absent) | 11 |
| FM6 | List of excipients not provided. | 8 |
| FM7 | The tablets/capsules not uniform in shape/size/Surface coatings/Texture/Scoring/Odour. Presence of breaks, cracks, splits, or pinholes; Tablets/capsules not free of embedded surface spots and foreign particle contamination | 12 |
| FM8 | Presence of empty tablets/capsules in the package | 13 |
| FM9 | Tablets not disintegrating. | 1 |
| FM10 | Identification: absence of Artemether in the dosage form | 4 |
| FM11 | Identification: absence of Lumefantrine in the dosage form | 5 |
| FM12 | Percentage label claim_Artemether content outside the allowed range (90-110%). Not more than 110%, and not less than 90% of artemether | 2 |
| FM13 | Percentage label claim_Lumefantrine content outside the allowed range (90-110%). Not more than 110%, and not less than 90% of Lumefantrine | 3 |
| FM14 | Tablet friability | 15 |
| FM15 | Tablets not uniform in weight | 14 |

The closeness coefficient provided the basis of ranking of all the failure modes (Table 10). FM9 recorded the lowest CCI signifying its closest proximity to the negative ideal solution, and thus the highest risk of all the analysed failure modes. FM9 relates to failure in the ACT tablets to disintegrate based on USP 41-NF36 in vitro disintegration monograph. Although the historic quality control data indicated that the ACT tablets were immediate-release, some of the dosage units failed to disintegrate even after 1 hour signifying critical quality failure.

Similarly, FM12 and FM13 recorded the second and third closest distances to the negative ideal solution. These failure modes relate to the non-conformances in percentage label claims of artemether and lumefantrine, respectively. The pharmacopoeia specified the acceptable label claims to be not less than 90% and not more than 110% of either of the active agents. However, the historic data indicate that both artemether and lumefantrine in the ACT tablets fell below the acceptable official threshold.

Concerning proximity to fuzzy positive ideal solution, FM14 recorded the highest closeness coefficient. The historic data indicated that the ACT tablets were mechanically strong as they were characterized by friability values within the pharmacopoeial acceptable limits. FM15 and FM8 were the second and third closest to the positive ideal solution, respectively.

Although, this novel approach is an enhanced adaptation of the tFMEA, it is not without inherent limitations. There are several variants of AHP for according criteria weights. This may likely affect the order of risk factor importance priorities, which may affect the categorization of the variables as beneficial and non-beneficial risk variables. We therefore recommend further studies to corroborate the current findings and also explore other variants of fuzzy multicriteria decision analysis in quality risk analysis of ACTs.

## Conclusion

FMEA is a crucial proactive quality risk analysis tool in lifecycle management of Artemisinin-based Combination Therapies (ACTs) to assure quality and safety status. The tradition FMEA methodology has critical limitations that compromises the reliability of risk prioritization. The advanced hybrid fuzzy FMEA methodologies presented in this paper is consistent with the current regulatory paradigm shifts from “empirical thinking” to “scientific thinking”, as contextualized in the ICH Q9 (Quality Risk Management) and Q10 (Pharmaceutical Quality Systems) guidelines.

The novel fuzzy FMEA model captures the diverse subjectivities of the cross functional team by fuzzification of the linguistic and crips values to fuzzy numbers and membership functions. Secondly, the analytical hierarchy process provided a reliable computational way of weighting the risk-determining variables. The fuzzy TOPSIS provides an easily configurable ranking of the alternatives (failure modes) on the basis of their Euclidean distances from positive and negative solutions.

This study targets to strengthen the capacity and scientific quality of quality risk analysis of ACTs by all stakeholders involved risk mitigation and containment strategies particularly in LIMCs where both malaria and substandard antimalarials are endemic.

## Data Availability

All data produced are available online via the link: https://data.mendeley.com/preview/hg7z83brcv?a=d8f53913-d5a9-4a49-8964-.

## Acknowledgement

We sincerely appreciate the funding received through the National Research Fund of the Tertiary Education Trust Fund Grant Number TETF/DR&D/CE/NRF/CC/20/VOL 1.

